# Software-defined Radar for MRI Motion Correction: A versatile, vendor-independent Platform

**DOI:** 10.64898/2026.05.16.26351399

**Authors:** Christoph Maier, Eddy Solomon, George Verghese, Hersh Chandarana, Kai Tobias Block, Leeor Alon

## Abstract

**Purpose:** To develop and evaluate a flexible, software-defined radar platform for contactless, vendor-independent motion detection and correction in MRI.

**Methods:** A continuous-wave (CW) Doppler radar was implemented using a software-defined radio and the open-source GNU Radio framework. The system was deployed inside a 1.5T MRI scanner and synchronized with MRI acquisitions. We evaluated the performance in a custom-developed internal motion phantom and in healthy volunteers to track respiration and bulk motion. The radar-derived signal was validated against cine MRI and used to demonstrate both retrospective and prospective motion management techniques in phantom and in healthy volunteers.

**Results:** The radar provided robust motion signals that correlated strongly with image-based ground truth signals in both phantom and volunteer experiments. Signal characteristics were found to be frequency-dependent, enabling optimization for different motion regimes. Retrospective correction of free-breathing abdominal data using the radar signal effectively suppressed respiratory artifacts, achieving image quality comparable to a self-gating approach. Prospective triggering successfully reduced motion artifacts in the phantom study. The system also reliably detected sporadic events such as swallowing during neck imaging.

**Conclusion:** Software-defined radar was demonstrated to be an effective platform for both prospective and retrospective motion correction. Its independence from the MRI system, ultra-wide band capabilities, and body-region versatility enable the adaptation of the technique for a wide range of imaging applications and protocols.

## Introduction

MRI has become an indispensable tool for diagnosing, staging, and treatment monitoring of disease in virtually every field of medicine (1, 2). However, compared with competing cross-sectional imaging modalities such as CT, a fundamental limitation of MRI is the slow nature of the acquisition process, rendering it susceptible to data corruption by motion. Various types of motion occurring during the acquisition can render MR images non-diagnostic and limit the applicability of MRI (3).

Motion in MRI presents as a multi-faceted problem and a wide range of strategies to prevent or correct motion artifacts has been developed (4, 5). Motion correction typically requires knowledge of the motion pattern. This information can be derived from MRI data itself using dedicated navigator echoes (6) or, for selected k-space trajectories, through self-gating (7). Alternatively, motion can be detected using external sensors placed within the MR environment, functioning independently of the imaging apparatus. While clinically established sensors have proven valuable, they have limitations. For example, the cumbersome setup of electrocardiography (ECG) increases total table time and is impacted by the magnetohydrodynamic effect, especially at high field (8, 9). A broad range of advanced MRI-compatible sensors has been proposed (10). These sensors monitor the body through various physical phenomena, including acoustics (11), light (12), or electromagnetism to detect motion.

RF-based sensing is particularly attractive due to its contactless nature and has recently gained renewed interest. This broad category includes passive techniques exploiting motion-induced modulation of the thermal noise of MR receive coils (13) or of the reflected power in the transmit path (14), active devices that measure loading of a loop coil (15, 16), and the pilot tone technique, wherein a constant RF tone is generated near the Larmor frequency to enable recovery of motion-induced modulation using the MR receive chain (17–19).

Several groups have also proposed RF sensors that employ radar principles for motion detection, including continuous-wave (CW) radar (20), frequency-modulated CW (FMCW) radar in the mmWave range (21), or ultra-wideband impulse radar (22, 23). An advantage of radar-based approaches is their independence from the MRI RF subsystem, allowing flexible selection of the operating frequency. Outside of MRI, the use of radar for biomedical sensing is a well-established field of research (24, 25).

Traditionally, radar systems have been implemented using custom-developed hardware components, implicating that cardinal system parameters are fixed during the design process. In the broader engineering community, software-defined radio (SDR) has emerged as an alternative paradigm to the conventional way of building RF systems from discrete components or application-specific integrated circuits(26). While a dedicated hardware unit (also called SDR) is still required, most of the radio functionality is shifted to the software domain. This offers significant flexibility in system architecture and in operational parameters such as the carrier frequency, bandwidth, and gain. Typically, the control and signal processing logic of an SDR system is executed on a general-purpose computer. SDR is typically programmed using flowgraphs - visual representations of algorithms composed of interconnected blocks - to define the signal flow and processing through the transmit and/or receive paths. This software-driven approach enables easy prototyping, shortens development-evaluation cycles, and can even enable dynamic reconfiguration of the RF system at runtime, making it well-suited for adapting to varying patient anatomies and physiological conditions.

In this work, we present a software-defined radar platform for MR motion correction. As proof-of-concept, we implement a CW Doppler radar and evaluate its capability to detect internal motion in a phantom setup as well as respiratory and bulk motion in volunteer experiments. Moreover, we demonstrate the utility of the radar-derived motion signal for retrospective and prospective MR motion correction.

## Methods

### Radar system

A single input, single output CW Doppler radar (27) was implemented in this work. The two antennas (1x Tx, 1x Rx) were mounted in a bistatic configuration on a half-cylindrical scaffold. Antennas were mounted laterally for phantom experiments (Figure 2B) and anterolaterally for volunteer experiments (Figure 1C). In z-direction, the antennas were positioned close to the isocenter in all experiments. We utilized ultra-wideband antipodal Vivaldi antennas previously designed in-house for biomedical microwave applications (28). Antennas were connected to a networked SDR unit (USRP N310, Ettus Research) positioned outside of the magnet room.

**Figure 1.**
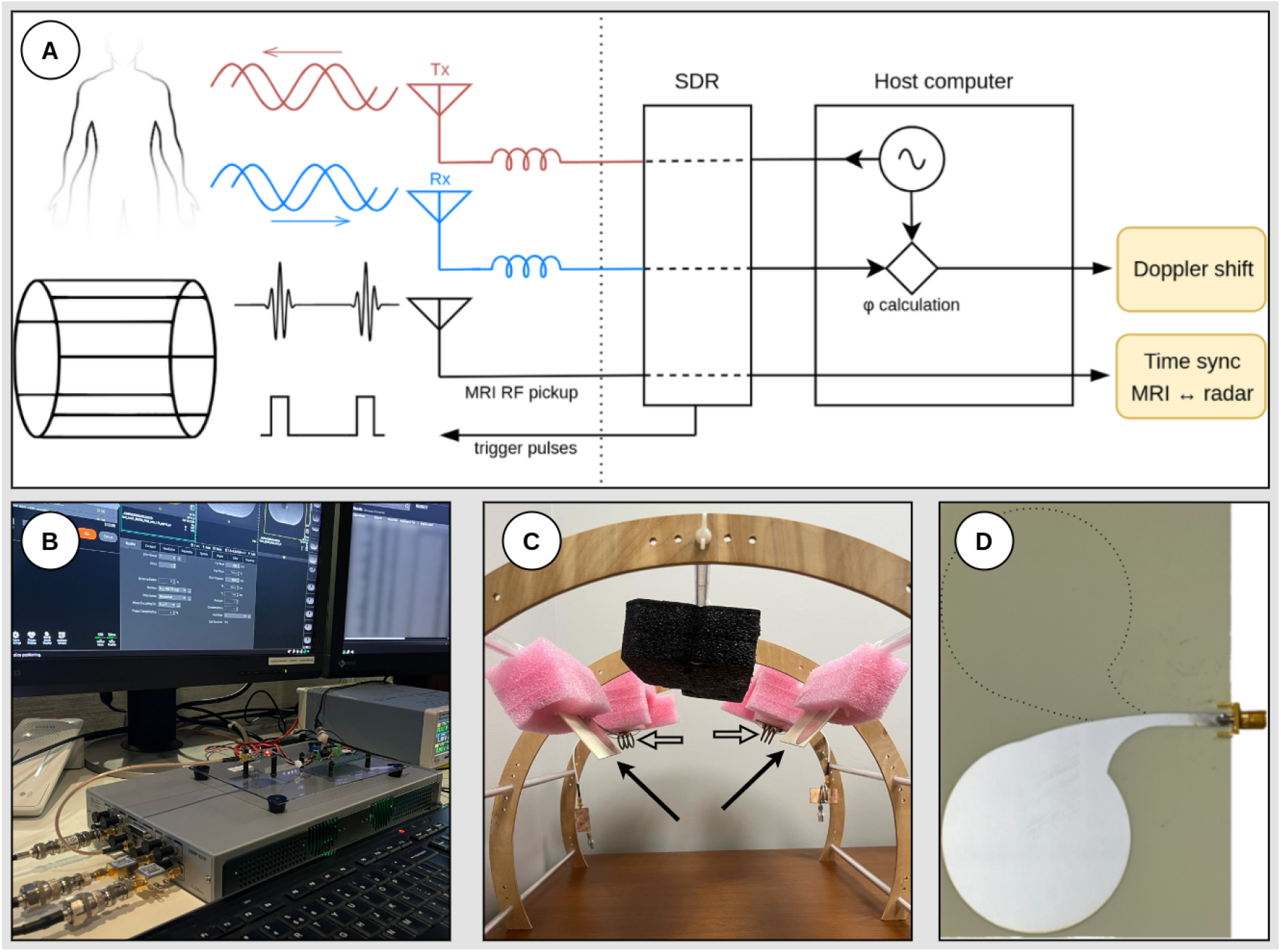
System overview. (A) Simplified scheme of signal paths for the radar (Tx/Rx), MRI RF pickup for time synchronization, and trigger pulses for prospective triggering. (B) Photograph of the SDR hardware in the control room. (C) Close-up of the scaffold holding antennas (arrows) and cable traps (open arrows). (D) Close-up of an ultra-wideband antipodal Vivaldi antenna (dotted line indicates the outline of the conductor on the reverse side).

**Figure 2.**
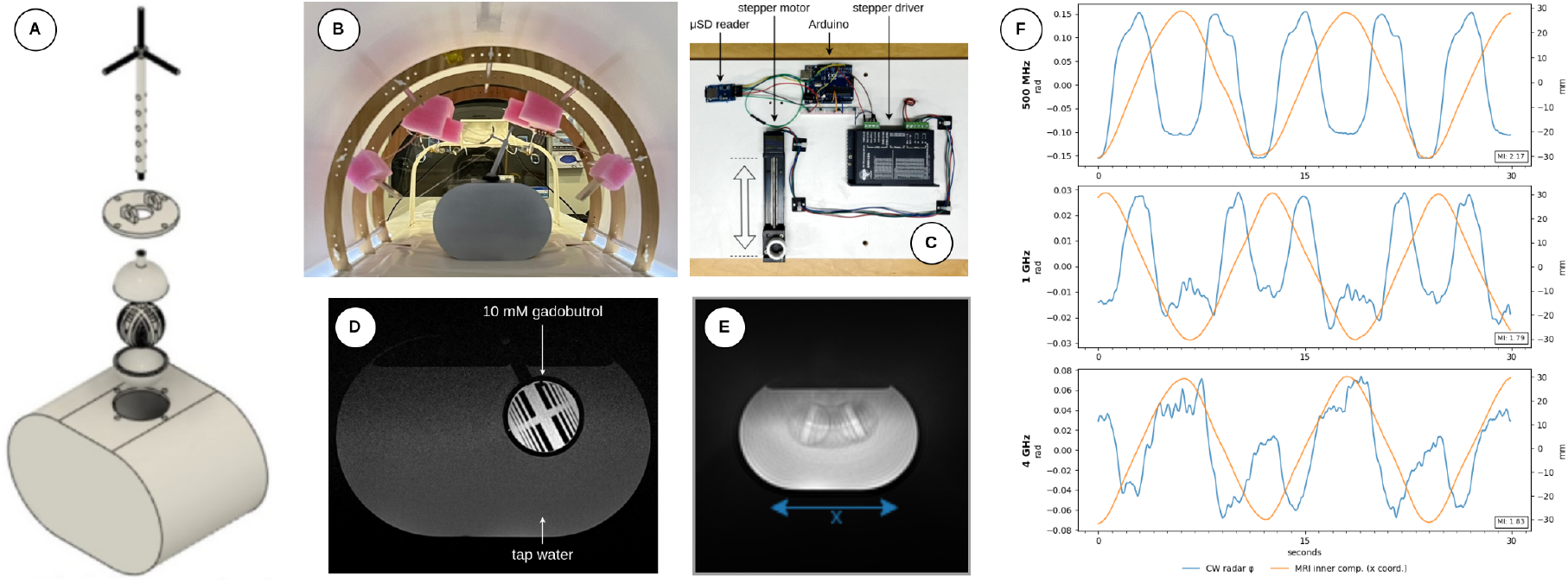
Detection of internal motion in a phantom setup. (A, B) CAD drawing and photograph of a 3D-printed phantom simulating internal body motion. A water tank houses the inner compartment, which is mounted on a rod. (C) The motor assembly was located outside the magnet room. A long string connects the rod to the linear motion stage, such that actuating the stepper motor results in pendular motion of the inner compartment. (D) T1w TSE image illustrating the internal structure. The hollow spaces between the printed line pairs in the inner sphere were filled with gadobutrol. (E) A time-averaged projection demonstrates the range of motion. (F) The phase shift between transmitted and received tones at baseband (orange line) is compared to the x-position of the internal compartment as determined from cine MRI (blue). The CW radar detects internal motion on all evaluated carrier frequencies, but the lowest frequency exhibits higher mutual information (MI) with the ground truth and lower noise, most likely due to electromagnetic penetration characteristics. Note that the phase shift alone does not map unambiguously to the internal compartment’s position in all motion states.

Considering the exposure to the transmit fields of the birdcage coil, common mode currents on the shields of the coaxial cables were suppressed with two tank traps on both the Tx and Rx paths (Figure 1C). Additionally, ferrite cores were attached to the cables outside of the magnet room (29). Band-stop filters (ZX75BS-5468-S+, Mini-Circuits) protected the SDR against differential mode currents.

MRI RF pulses were recorded via a small loop antenna taped to the wall of the magnet room (30). This approach was facilitated by the specific SDR model employed, which is equipped with two RF front ends. These front ends operate on independent local oscillator frequencies, allowing one front end to be used for the radar acquisitions, while the second frontend was tuned close to the scanner’s center frequency (63.65 MHz) for synchronization of the SDR system with the MRI system. This architecture was useful since it was intended to build a motion sensing system that is vendor independent.

The CW Doppler radar system was operated using GNU Radio (v3.10.10.0), a free and open-source SDR software framework (31, 32). This platform features a visual development environment for constructing signal processing applications as flowgraphs, where processing blocks are interconnected to define the path of a data stream. A library of highly optimized implementations of common signal processing kernels (33) is available, combined with the possibility to implement custom logic. This extensibility was expedient for implementing the flowgraph for prospective triggering (see below), which required custom Python blocks to ensure stable data streaming. Based on published latency measurements for older GNU Radio/USRP platforms(34), we adopted a design assumption of at most 10 ms end-to-end latency across transmission, reception, and host-side processing in the CW Doppler radar system. This value was intended as a conservative upper estimate rather than a direct measurement of our setup.

As baseband signal, a 1 kHz complex sinusoid of constant amplitude was generated in the GNURadio flowgraph and streamed to the SDR over ethernet, where it was upconverted to the carrier frequency (evaluated frequency range: 500 MHz - 4 GHz). Conversely, the reflected wave was downconverted to the baseband and sampled in quadrature (typically at 125 MHz, the lowest available ADC clock speed), decimated by a factor of 1000 in the FPGA, and streamed back to the host computer. Real-time phase demodulation was implemented in GNURadio, i.e. the complex conjugate of the received baseband signal was multiplied with the transmitted reference tone, followed by complex argument calculation, yielding the phase difference(32). The phase difference was then used for the various motion-correction strategies. The baseline phase difference has an initial offset, influenced by the distance between transmitting and receiving antennas, and signal propagation in the coaxial cables.

All signal data were recorded using the community-defined SigMF standard(35), facilitating offline analyses through a well-defined metadata schema for parameters such as sampling rate or carrier frequency.

### Signal processing

For retrospective analyses, the phase shift was re-calculated offline after band-pass filtering the recorded raw baseband stream to suppress noise induced by the MRI RF pulses. Next, the phase shift was further decimated (target rate 50 Hz) and smoothed with a Savitzky–Golay filter (window length of 1 s, 5th order polynomial)(36). Time synchronization between the MRI and the standalone radar system was achieved through the MRI RF pulses recorded in parallel with the radar signals. For this purpose, the time points of all MRI RF pulses were identified as the midpoints of contiguous “on” segments in a binary trace obtained by amplitude-thresholding of the recorded pickup signal. Depending on the respective pulse sequence and acquisition parameters, time synchronization was improved by accounting for dummy pulses, fat-saturation pulses, and, in the case of turbo spin echo (TSE), differentiating between excitation and refocusing pulses.

### Motion phantom

We modified a previously described 3D-printed phantom to emulate internal body motion (37), see Figure 2 A-E. A water-filled outer compartment encloses an inner compartment. This inner compartment is a hollow sphere containing 3D-printed line pairs with widths between 0.25 and 4 mm and 10 mM of gadobutrol. The sphere was secured to a rod protruding from the outer compartment, creating a pendulum. A string attached to the rod was pulled by a stepper motor. To ensure reproducible motion patterns, the motor was driven by a microcontroller board (Arduino Uno Rev 3, Arduino S.r.l.). This setup allowed the placement of all ferromagnetic components safely outside of the ACR zone IV. To increase the range of motion, a rubber band pulling in the opposite direction was also attached to the rod, as the string cannot be pushed by the stepper motor.

### Experiments

The radar system was evaluated using a whole-body 1.5T scanner (MAGNETOM Sola, Siemens Healthineers; version XA51A) in a phantom and in healthy volunteers, as described in detail below. All studies involving human subjects were approved by the institutional review board.

#### Cine MRI experiments

Radar and single-slice 2D cine MRI were acquired simultaneously to compare radar-derived motion signals against the image-based ground truth. In the phantom, single-slice measurements were acquired in the plane defined by the center of the outer container and the direction of motion along the phantom’s long axis. For MRI of the human chest, the sagittal plane through the apex of the hemidiaphragm was imaged. For neck experiments, an oblique slice aligning with and encompassing the true vocal cords was imaged. Depending on the desired contrast and flow sensitivity, either a golden-angle radial spoiled GRE sequence leveraging view-sharing and sliding-window reconstruction for high frame rates (11.8 fps) (38) or a Cartesian b-SSFP pulse sequence (4.2 fps) was employed. For some cine volunteer experiments, data from a respiratory sensor embedded in the spine coil of the MR system were also recorded (BioMatrix Spine32, Siemens). To ensure reliable measurements with this sensor, care was taken to position the chest according to the manufacturer’s instructions.

#### Motion correction for radial MRI

Radar data were acquired in parallel with a 3D stack-of-stars spoiled GRE acquisition of either the motion phantom (periodic motion pattern) or the human abdomen under free breathing. Each k-space line was sorted into discrete motion states according to the radar phase shift, followed by a motion-resolved reconstruction (39) using the BART toolbox (40). To compensate for the undersampling resulting from motion binning, a compressed-sensing approach (41) was used, applying a total-variation constraint along the motion dimension. The regularization coefficient was manually tuned for each imaged region (phantom vs. abdomen). For comparison, the identical reconstruction technique was separately applied to data binned using the self-gating signal from the center of k-space (42), and the full, unbinned dataset was reconstructed conventionally. Human datasets were further post-processed using the SimpleITK toolkit(43): N4 bias field correction was applied to correct for B1+ inhomogeneities and multi-planar reformation of the anisotropic volume was performed using B-spline resampling.

#### Motion correction for Cartesian MRI

The utility of the radar system for correcting aperiodic motion in Cartesian acquisitions was evaluated using an adaptive re-acquisition strategy (44, 45). Because of the significant implementation effort required for interfacing with the vendor-specific real-time acquisition control system, this strategy was retrospectively simulated as a proof of concept. To generate a data pool for this simulation, a 2D TSE sequence was acquired with full k-space sampling and a high number of signal averages (NEX = 10).

Two image reconstruction approaches were performed offline for comparison. First, an uncorrected reference image was generated from a single signal average. Second, a motion-corrected image was reconstructed from the same average. For the corrected reconstruction, motion-corrupted k-space lines were identified using the time-synchronized radar phase shift. Each corrupted line was then replaced with the first available uncorrupted line of the same slice and phase-encoding index drawn from the spare data pool created by the additional averages.

This methodology was tested in two scenarios: 1) The motion phantom was programmed to undergo pseudo-random sporadic movements, wherein the inner compartment would rest at the center position but jiggle back and forth sporadically. The number of jiggle excursions, jiggle amplitude, and the delay until the next jiggle followed a pseudo-random pattern. 2) A healthy volunteer performed instructed jaw-opening and -closing maneuvers during a coronal 2D TSE acquisition of the midface.

### Prospective triggering

To evaluate the system’s capability for prospective motion management, the radar system was used to trigger a T2-weighted BLADE sequence during a phantom experiment. The radar signal was processed in real-time within the GNU Radio flowgraph to identify a predefined state of the phantom’s periodic motion cycle. Whenever the system detected this motion state, bursts of on/off pulses were generated and transmitted through an unused channel on the radio frontend tuned for MRI RF pickup. This on-off keyed RF carrier signal was demodulated into a 5V transistor-transistor-logic (TTL) signal using an external circuit composed of a logarithmic RF power detector and a voltage comparator. The resulting TTL pulse was supplied to the MRI scanner’s external trigger input, triggering the BLADE sequence to acquire k-space data only during the desired motion state. An untriggered acquisition using the same motion pattern was acquired for comparison.

### Data analysis

For phantom experiments, the position of the spherical inner compartment was tracked by applying a Canny edge detector followed by a Hough circle transform to each cine frame, taking the known diameter into account. This circle-detection approach was facilitated by prescribing the imaging plane slightly off-center, thereby excluding the stalk of the inner compartment. Because the pendulum motion also caused flow-related signal alterations in the surrounding water on spoiled GRE, cine frames were pre-processed with local histogram equalization for increased robustness of the segmentations.

To establish ground truth respiratory motion in human experiments, diaphragmatic breathing was defined as head-foot translation of the diaphragm and costal breathing was defined as antero-posterior translation of the anterior chest wall, as observed in a sagittal cine acquisition of the right hemithorax. For diaphragmatic motion, a vertical pixel column was extracted near the apex of the hemidiaphragm. The high-contrast interface between the lung and liver formed the dominant rising edge within the resulting 1D signal intensity profile. The position of this edge was localized by convolving the profile with a step function, followed by an argmax operation. For costal motion, the same process was applied to a horizontal pixel row in a suitable location, tracking the boundary between external air and subcutaneous fat.

To compare the radar data with the movement of individual organs, the organs were tracked frame-by-frame using the general-purpose image segmentation model SAM 2 (46), following pre-processing with contrast-limited adaptive histogram equalization (47) and manual annotation of seed points on a small number of frames. The concordance between various signals was assessed using Pearson’s coefficient of correlation (CC) and by mutual information (MI).

Image quality in motion corrected radial MRI was assessed using region-based histogram analysis of the signal intensity for phantom experiments and by quantifying 20% to 80% signal-intensity distance of the rising edge at the lung-liver boundary for human experiments.

All manual annotations and all automated segmentation were created and verified by a board-certified radiologist, respectively.

## Results

### Phantom experiments

The in-bore motion-sensing capabilities of the software-defined CW radar system were first evaluated in a phantom setup (Figure 2A-E). For a set of experiments in which cine MRI and radar were acquired simultaneously while the inner compartment of the phantom underwent a periodic motion pattern, Figure 2F shows the ground truth position of the inner compartment plotted against the radar phase shift (both normalized). Using identical motion patterns, this experiment was repeated multiple times with the SDR tuned to carrier frequencies of 500 MHz, 1 GHz, and 4 GHz, respectively. The radar phase shift was sensitive to the positional changes on all evaluated frequencies, demonstrating the capability to sense internal motion. The waveform of the phase shift over time and the noise level were frequency-dependent, with 500 MHz yielding the most robust motion sensing in this setup (mutual information between the ground-truth x-position of the mobile sphere and the normalized radar phase shift at 500 MHz vs. 1 GHz vs. 4 GHz was 2.17, 1.79, and 1.83, respectively). With this 1 Tx, 1 Rx antenna configuration interrogating a reflector undergoing two-dimensional arc motion, correlating the phase shift directly to the displacement resulted in ambiguities for certain motion states.

In the same periodic motion pattern, retrospective motion correction of a 3D golden-angle stack-of-stars GRE acquisition was achieved by binning each k-space line to a motion state according to the radar phase shift (500 MHz carrier frequency). In the subset of motion bins where the absolute position could be unambiguously determined from the radar phase shift, the motion-resolved reconstruction resulted in a marked reduction of motion artifacts compared to a conventional (motion-averaged) reconstruction (Figure 3). Of note, the region-based histogram analysis was chosen because some motion bins do not resolve the position of the moving sphere unambiguously, impeding an accurate / fair segmentation of the moving object.

**Figure 3.**
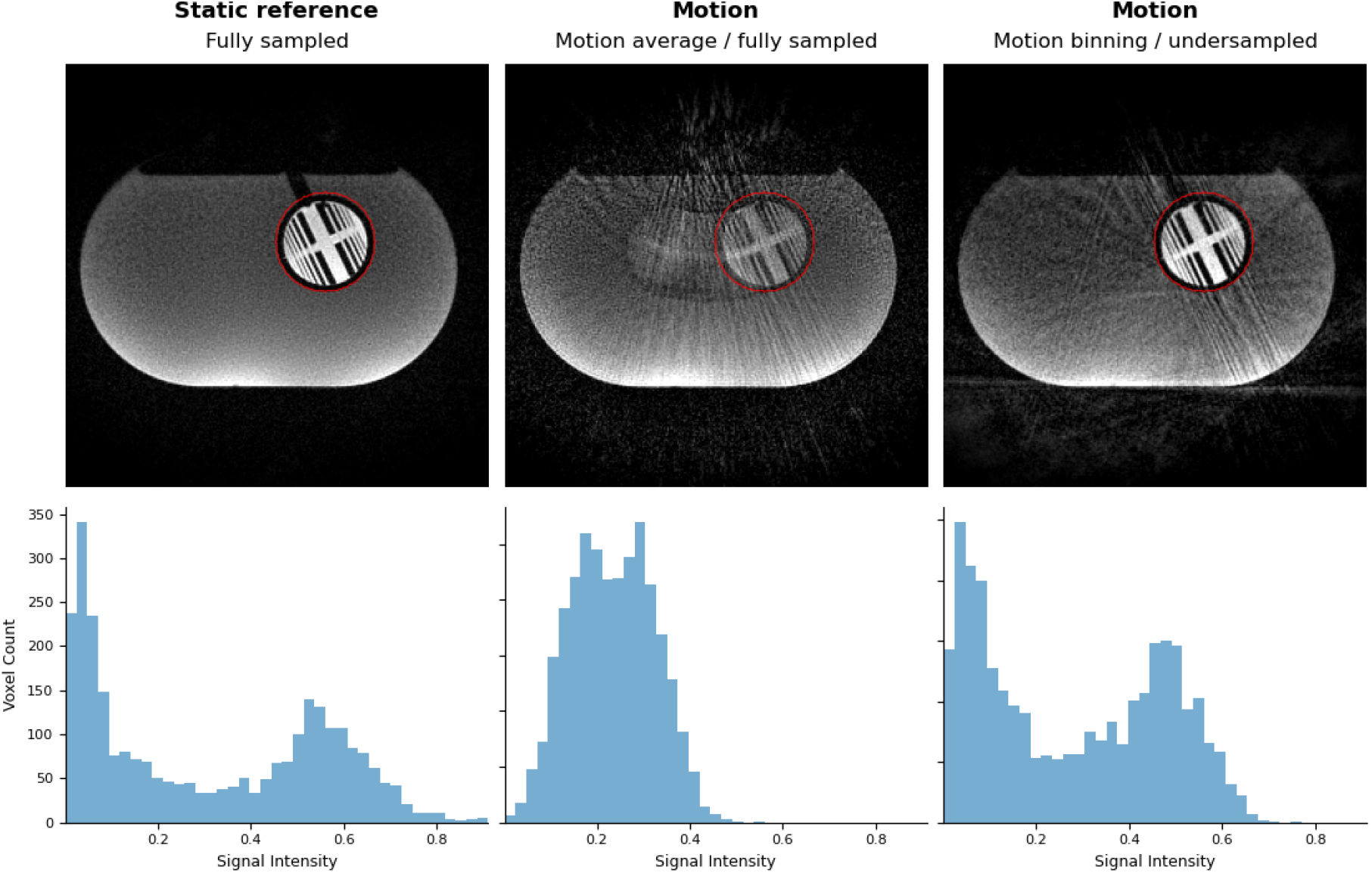
Retrospective motion correction in a phantom setup. **Upper row:** 3D stack-of-stars GRE data acquired with the phantom stationary (left). In a separate acquisition, the phantom was undergoing periodic motion while the radar system operated at a 500 MHz carrier frequency (center, right). For the motion-resolved volume, k-space data were retrospectively sorted into 6 discrete motion states according to the radar phase shift, followed by a compressed-sensing reconstruction with total-variation regularization along the motion dimension (right; best bin shown). Compared with the motion-averaged reconstruction (center), this approach successfully suppresses motion artifacts. **Lower row:** Histogram analysis of signal intensities within a stationary circular region targeting the resolution insert, annotated in red. Under static conditions (left), a distinct bimodal signal intensity distribution is observed, corresponding to the high-signal gadolinium-doped water and the signal-free 3D-printed line pairs. In the conventional motion-averaged reconstruction (center), the object’s continuous transit through the fixed ROI blurs the internal geometry, collapsing the bimodal distribution. The radar-based, motion-resolved reconstruction (right) enables the recovery of the internal contrast, demonstrating the preservation of spatial features despite the object’s motion.

In a complementary experiment, the utility of the radar system for prospective triggering was tested in the same phantom setup. The radar data were processed in real-time to trigger data acquisition for a T2w BLADE sequence. Compared to the untriggered acquisition, this method effectively suppressed motion artifacts (Figure S1).

### Respiratory motion

In the next phase of the study, the radar system was tested in healthy volunteers. Respiratory motion was selected as an initial test case due to well-established reference respiratory sensing methods and the ability of volunteers to control it for experimental purposes. Figure 4A shows the M-mode display of diaphragm motion overlaid with the radar phase shift, again revealing frequency-dependent results: At a 1 GHz carrier frequency, the phase shift closely tracked the ground truth when the volunteer was instructed to perform deep breathing. During shallow breathing, however, either noise or superimposed motion signals originating from other structures became noticeable. At 4 GHz, by contrast, a high-quality respiratory trace could be recovered even under shallow breathing. The radar phase shift (4 GHz) was compared to the respiratory signal provided by a commercially available coil-load respiratory sensor located within the spine coil assembly Figure 4B). While both sensors provided a usable respiratory surrogate, the radar phase shift exhibited superior accuracy in this experiment (MI: 1.52 vs. 0.91 for the radar and coil-load sensor, respectively). Furthermore, in an experiment employing a 3D stack-of-stars sequence, the radar phase shift (4 GHz) was in excellent agreement with the self-gating signal derived from the k-space center (Pearson’s CC: 0.99; Figure 4B).

**Figure 4.**
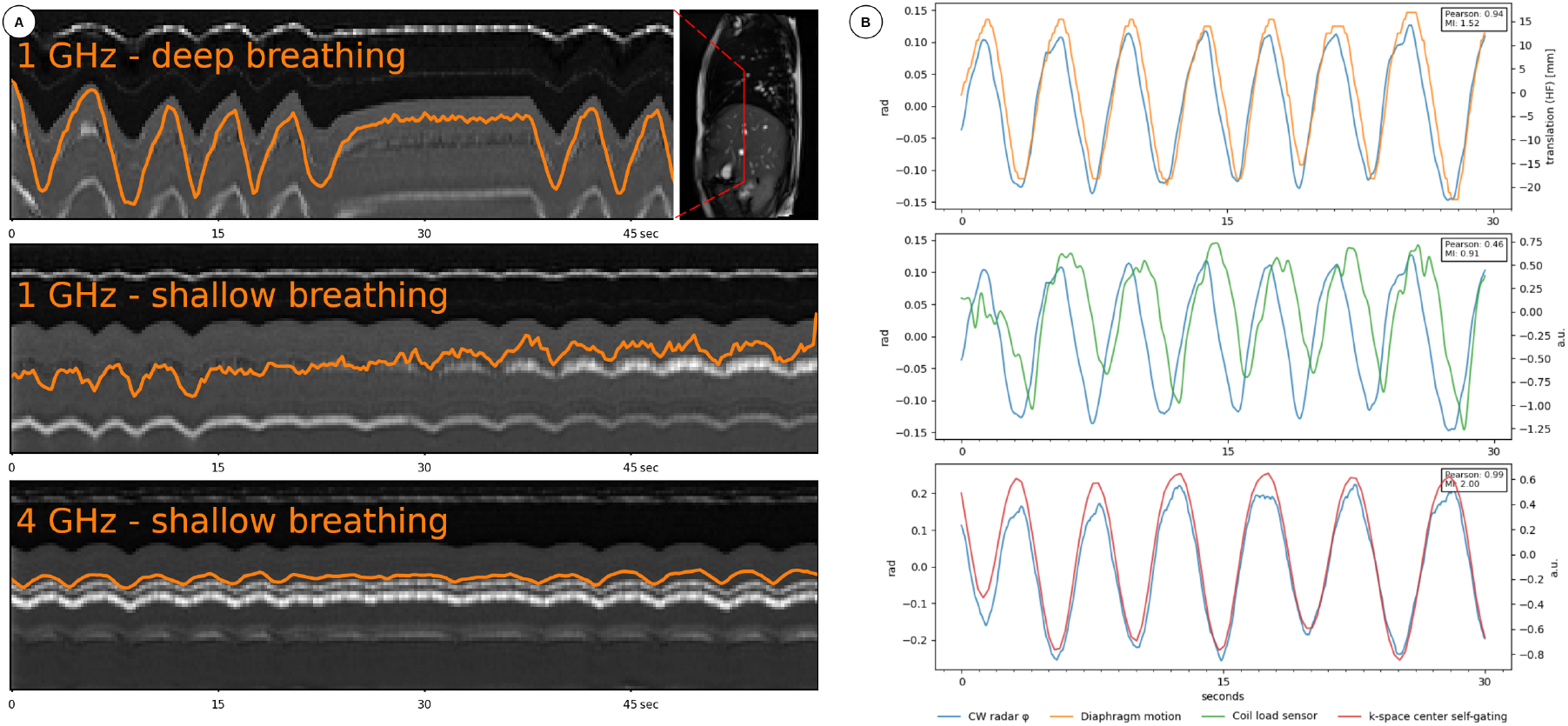
Respiratory motion sensing. (A) Radar data and cine b-SSFP MRI of the right torso were acquired simultaneously. The cine MRI data is plotted over time, and the radar phase shift is overlaid in orange. Under deep breathing, the radar phase shift closely follows diaphragm motion at a 1 GHz carrier frequency, but noise became noticeable during shallow breathing, possibly representing the superposition of internal motion such as heartbeats or peristalsis. Increasing the carrier frequency to 4 GHz increases SNR for respiratory tracking, such that a robust trace can be recovered even under shallow breathing. Note: For visualization purposes, the scale of the radar traces was individually adapted to the cine MRI. (B) Using data from a comparable experiment in a different volunteer, the radar-derived respiratory curve (at a carrier frequency of 4 GHz) was numerically compared to diaphragm motion in the head-foot direction (top row) as well as to the signal provided by a coil-load respiratory sensor embedded in the MRI system (middle row). The radar phase shift showed excellent agreement to the image-based reference. A moderate phase shift between radar phase shift and coil load sensor is evident, possibly attributable to inaccuracies in synchronizing the proprietary sensor data. As a further point of comparison, the radar phase shift is also highly correlated to the self-gating signal derived from the k-space center of a free-breathing 3D stack-of-stars acquisition (bottom row).

To further characterize the respiratory sensing performance, a volunteer was instructed to modulate the tidal volume within the duration of one cine MRI acquisition (Figure S2). While the respiratory-induced radar phase shift closely aligned with the position of both the diaphragm and the anterior chest wall in terms of periodicity and directionality, the wide dynamic range in this experiment revealed a better correlation with the anterior chest wall than with the diaphragm, suggesting that reflections from this structure were the dominant source of the radar-derived motion signal under these conditions (carrier frequency 1 GHz).

Finally, the radar phase shift was compared to parenchymal organ motion, using the kidney as an example (Figure 5). The radar phase shift was in excellent agreement with the organ position, further supporting its suitability as a surrogate for respiratory motion.

**Figure 5.**
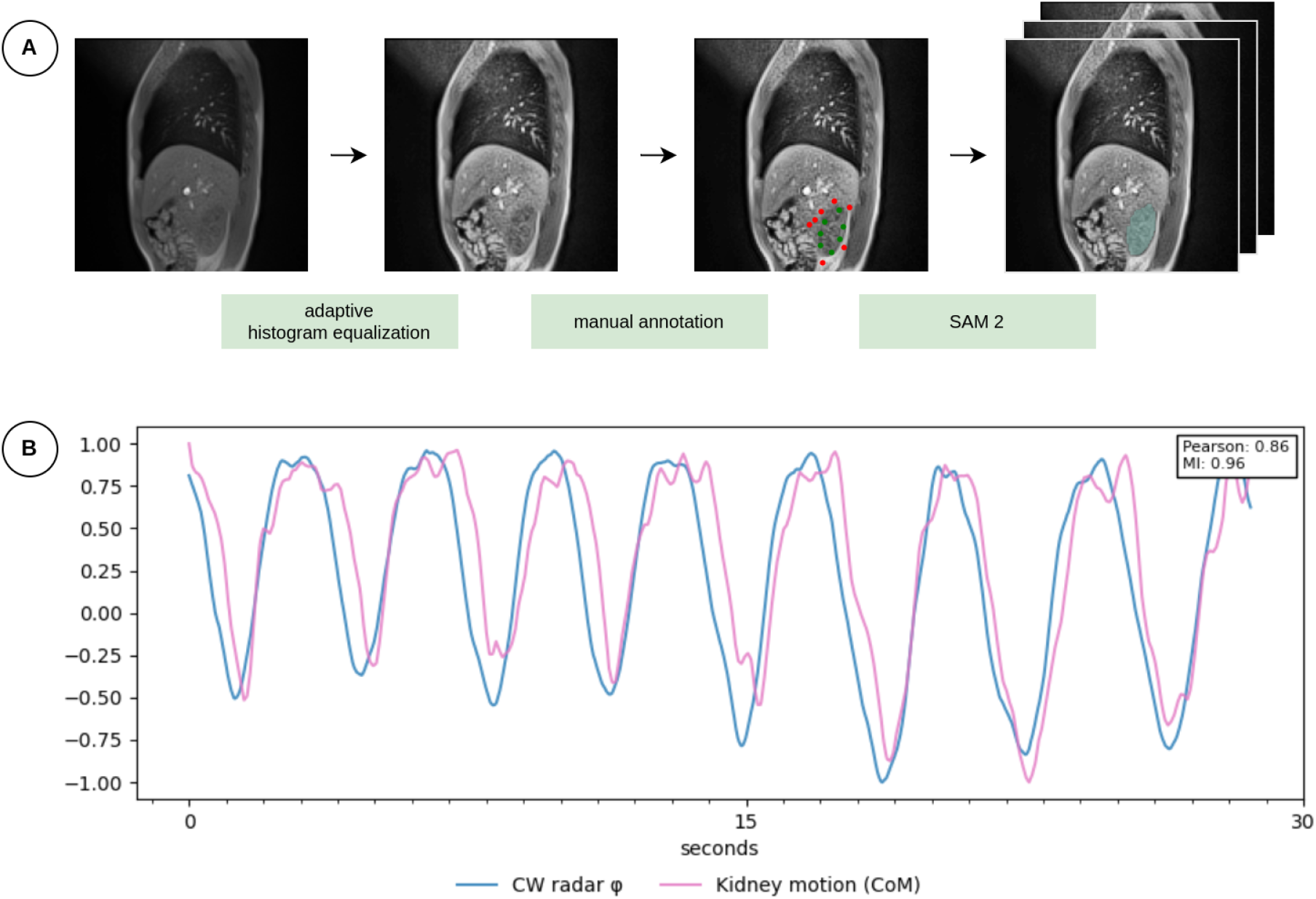
Correlation with organ motion aided by deep learning, using the kidney as an example. (A) Scheme of the segmentation process: Because the T1w GRE cine MRI was acquired without gadolinium administration in the healthy volunteers, contrast at the liver/kidney interface is low. Adaptive histogram equalization boosts contrast at this edge, aiding automatic segmentation. Positive (green) and negative (red) seed points were manually annotated on a few (usually one or two) frames. SAM 2, a general-purpose image segmentation model, was prompted to generate an organ mask from the seed points and to propagate this mask throughout all frames. 2D organ position was defined as the center of mass of these masks. The fidelity of the segmentation is demonstrated in Video S3. (B) Results from a representative experiment demonstrate excellent agreement between the Doppler radar phase shift and the magnitude of the position vector (both signals normalized).

To demonstrate practical utility for clinical MRI, the radar-based respiratory signal at a 4 GHz carrier frequency was used for a motion-resolved image reconstruction of free-breathing abdominal MRI. Similar to the motion-resolved phantom imaging regime described above, k-space data from a 3D golden-angle stack-of-stars sequence was binned into discrete respiratory states based on the phase shift, followed by a compressed sensing reconstruction regularized over the motion dimension. The radar-based retrospective motion correction markedly reduced motion blurring at a level comparable to a motion-resolved reconstruction using data binned according to the self-gating signal, which served as a reference method in this acquisition scheme (Figure 6). Both self-gating and radar resulted in reduced blurring of the diaphragm (width of the rising edge at the lung-liver boundary (20%–80% distance) for motion-average vs self-gating vs radar: 5.2 px, 1.7 px, and 1.5 px, respectively, assessing the respective best bin).

**Figure 6.**
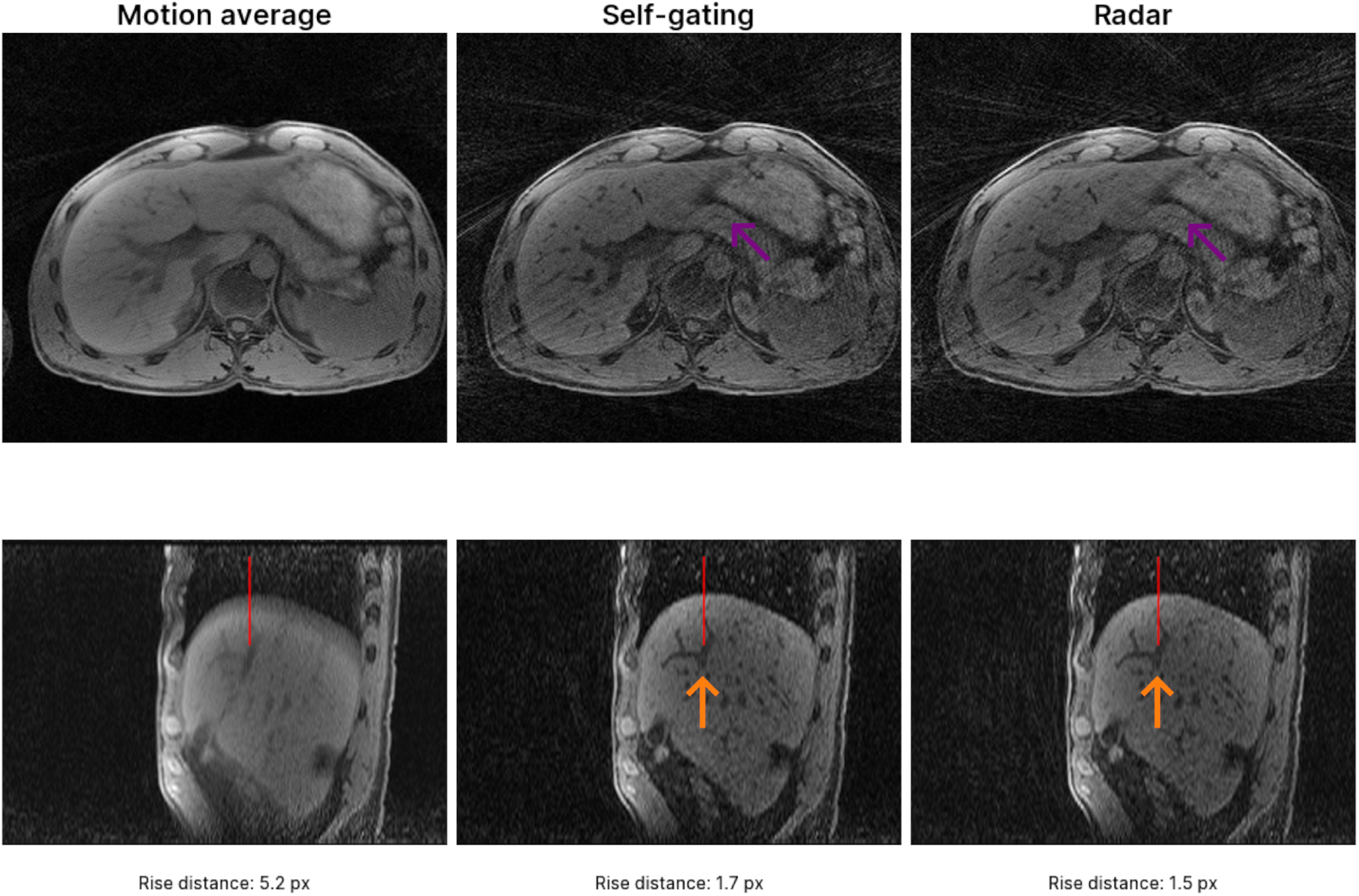
Respiratory binning. 3D stack-of-stars GRE data of the upper abdomen was acquired during free breathing. K-space lines were sorted into 5 respiratory bins according to the radar phase shift (at a carrier frequency of 4 GHz). For comparison, the same reconstruction was also performed using self-gating. Compared to the motion-averaged reconstruction, both methods suppress respiratory artifacts, resulting in a sharper depiction of structures such as the main pancreatic duct (purple arrows), intrahepatic branches of the portal vein system (orange arrows), or the diaphragm. Image sharpness was assessed by extracting 1D line profiles across the diaphragm boundary (indicated by red lines). Edge sharpness was quantified by calculating the 20% to 80% rise distance of the normalized signal intensity profile, utilizing high-density linear interpolation for sub-pixel accuracy. The results demonstrate a substantial reduction in edge blur in the motion-resolved reconstructions (best bin shown).

### Motion in the head & neck

To explore the motion-sensing potential of the CW radar system beyond the chest, the software-defined radar was operated concurrently with cine MRI of the head and neck, with the antennas pointing at the neck (Figure 7). The waveform of the radar phase shift was morphologically congruent with the signal from the vendor’s sensor embedded in the spine coil, indicating that the radar system could serve as a respiratory sensor in this position as well. A moderate temporal lag between the two signals was observed. Moreover, controlled swallowing events could be detected using the radar system, manifesting as a sharp spike in the phase shift (t=5 s, Figure 7E). Reliable mapping of this radar manifestation to swallowing events was enabled by signal enhancement of the paralaryngeal soft tissues in the time-synchronized spoiled GRE cine sequence, consistent with through-plane motion (Video S4).

**Figure 7.**
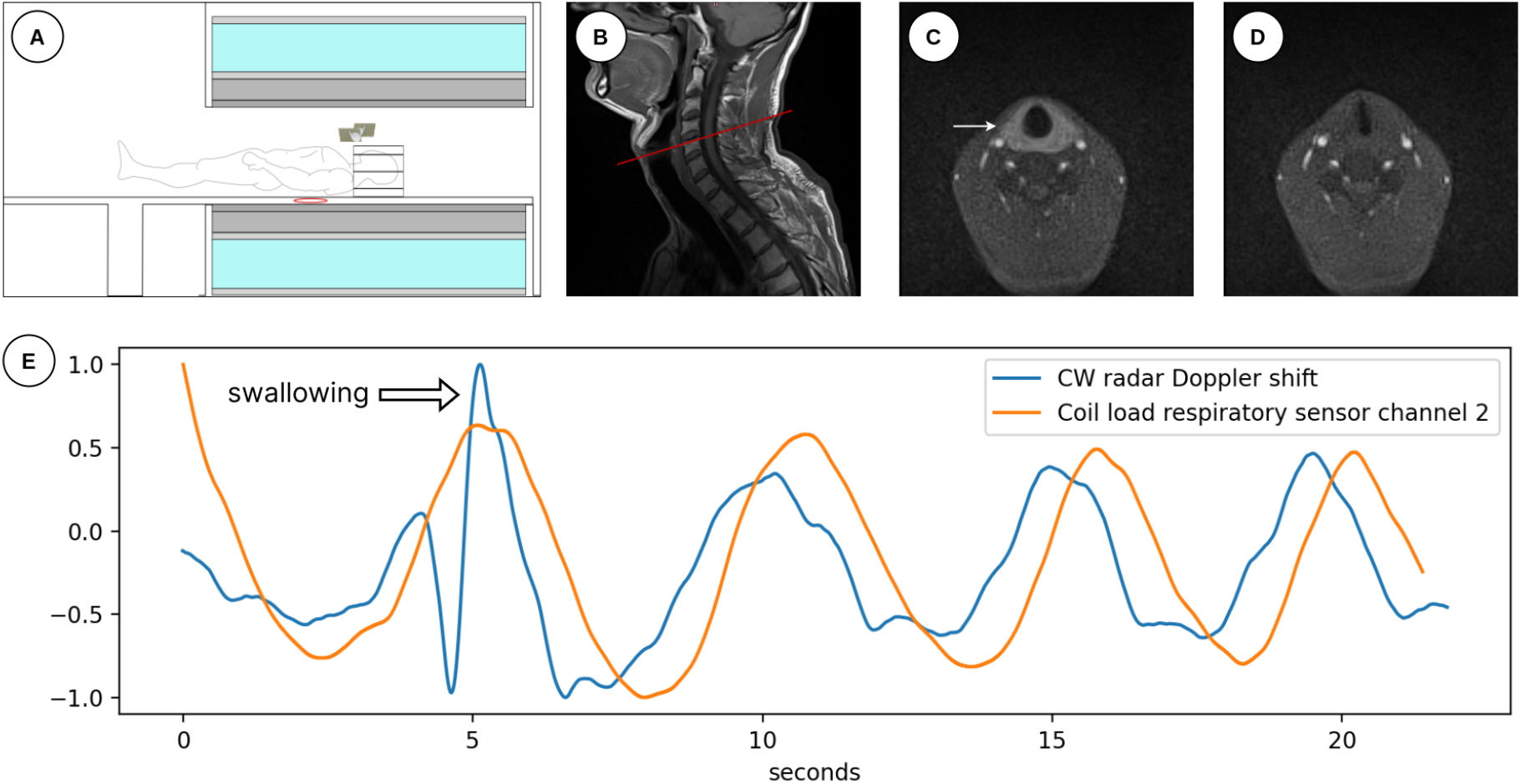
Motion sensing in the head and neck. (A) Scheme of the experimental setup. The radar Tx and Rx antennas were positioned just above the head/neck receive coil in a bistatic configuration, pointing at the soft tissues of the neck, whereas the coil-load respiratory sensor is integrated into the spine coil of the MRI system (symbolized as a red loop). (B, C) A cine radial GRE acquisition was prescribed in-plane with the true vocal cords. This serves as ground truth for swallowing events, as the associated through-plane motion leads to a pronounced signal enhancement of the pharyngeal and laryngeal soft tissue sleeve (arrow). (D) Cine frame from the same acquisition under resting conditions for comparison. (E) Comparison of motion signals: the CW radar phase shift (at a carrier frequency of 4 GHz) and the trace from the coil-load respiratory sensor, each normalized. The two signals for respiration are morphologically congruent with a constant temporal lag, while the CW radar additionally captures a swallowing event at t=5 s, readily identifiable by a sharp spike in phase difference.

Finally, the potential for radar-based correction of sporadic motion events was investigated. We utilized a retrospective simulation of an adaptive reacquisition strategy, where radar-identified motion-corrupted k-space lines were replaced with intact data from a spare pool (see Methods). This methodological framework was developed using the motion phantom, reprogrammed for aperiodic motion (Figure 8, upper row), and subsequently applied to a volunteer performing jaw opening/closing maneuvers during a coronal 2D TSE acquisition of the midface (Figure 8, lower row). In both cases, the radar-based correction markedly improved image quality by reducing ghosting artifacts.

**Figure 8.**
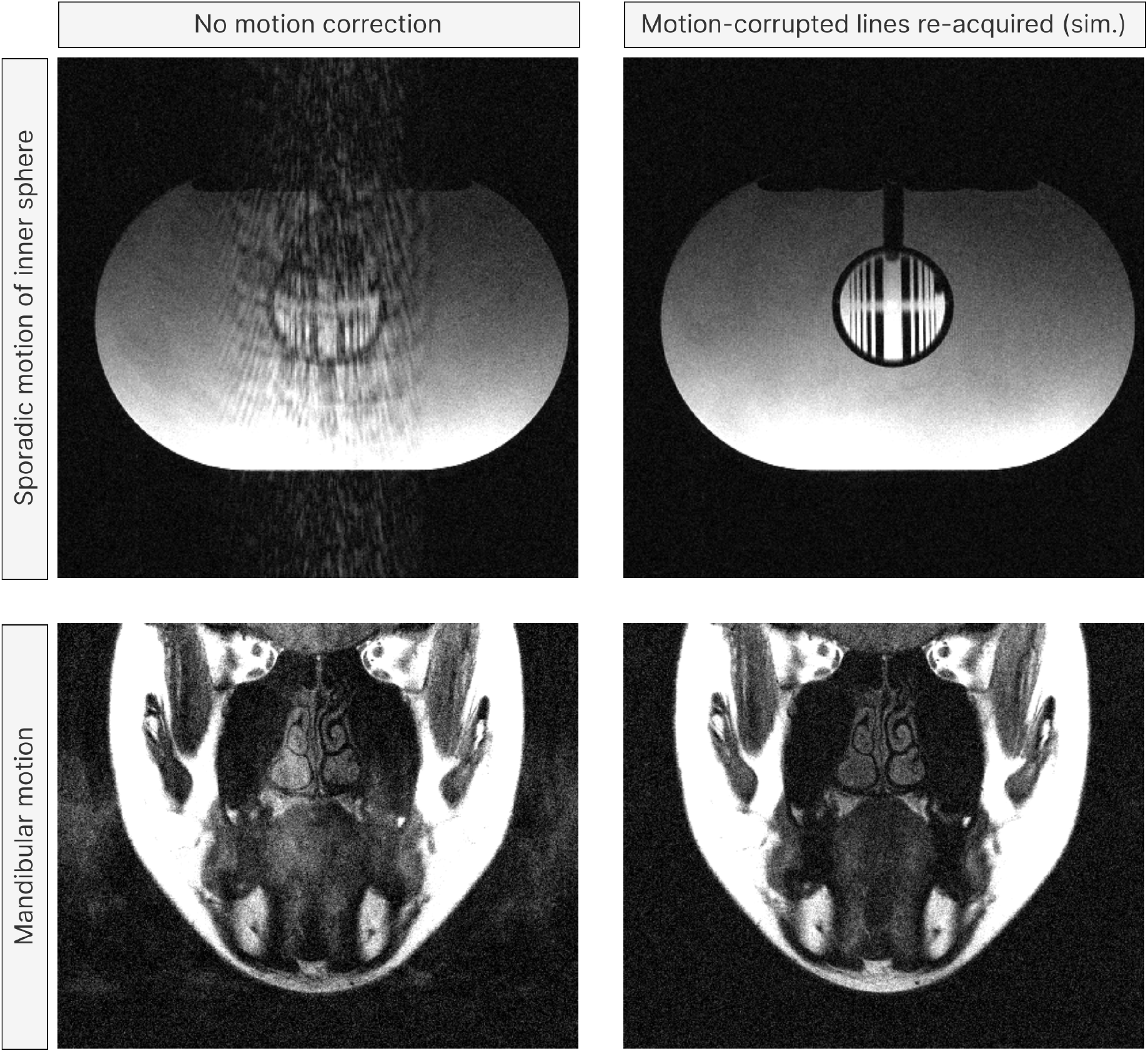
Simulated adaptive reacquisition for Cartesian data. Utility of the radar system for motion correction of Cartesian data was evaluated in two experimental setups using a 2D TSE sequence with sporadic (aperiodic) motion. Adaptive reacquisition was retrospectively simulated by acquiring fully sampled k-space with an abundance of signal averages (NEX = 10) and using spare, uncorrupted lines to replace motion-corrupted data, as described in Methods. Images were reconstructed offline either without motion correction (left column) or with radar-guided replacement of motion-corrupted phase-encoding steps (right column) Upper row: 2D TSE of the motion phantom (TR/TE = 680/16 ms) with pseudo-random jiggle motions of the inner compartment. Lower row: T1w coronal 2D TSE of the midface (TR/TE = 680/19 ms). The volunteer was instructed to perform jaw opening/ closing maneuvers. In both scenarios, the radar-based replacement of motion-corrupted data markedly reduces ghosting artifacts.

## Discussion

Here, we described a novel, vendor-independent radar platform for motion detection and motion correction that can operate across a broad frequency range and integrates into the MR environment. As a foundational proof-of-concept, we implemented a CW radar regime, which reliably detected motion signals across a range of scenarios. Key findings include the sensing of internal motion in a phantom and accurate respiratory tracking. Our data on respiration aligns with a recent study by Lee et al. (20). We extend these results by demonstrating the frequency dependency of the motion signal, detecting sporadic motion events in the head and neck, and showcasing the utility of the radar signal for both retrospective and prospective motion management techniques in radial and Cartesian MRI acquisitions.

Considering that a wide range of approaches for motion detection in MRI have already been developed, the radar technique needs to be compared with existing solutions.

Motion-sensing techniques based on the MR signal itself have the undeniable advantage that no additional hardware is required, effectively offering software-defined motion detection. Self-gating is an elegant technique for the subset of trajectories that continuously sample the k-space center, but it often fails for acquisitions with concurrent changes of the k-space signal intensity, such as during the administration of contrast agents or after generation of preparation pulses (48). Dedicated navigators overcome this limitation, but they must be synchronized with the host sequence, may increase acquisition time or disturb the magnetization state, and can be vulnerable to scanner instabilities.

In contrast to established external devices such as ECG or respiratory bellows, contactless sensors are strongly preferable from the perspectives of workflow, patient comfort, and hospital hygiene.

Of particular interest is the comparison of the software-defined radar with other contactless, RF-based motion sensing approaches. The pilot tone concept offers two distinct advantages: the required hardware is easier to integrate with the scanner than a standalone radar solution, and since the receive coil arrays on modern scanners typically offer a high channel count, the pilot tone yields a high dimensionality in the motion signal. However, the pilot tone is inherently linked to the Larmor frequency. Since the propagation of electromagnetic waves is frequency-dependent, this means it cannot tap into the full potential of RF motion sensing. The related Beat Pilot Tone technique avoids this limitation by transmitting two out-of-band tones that are mixed into a single, detectable signal within the MR receive band (19). However, Beat Pilot Tone also imposes additional hardware constraints, either in the form of vendor-specific non-linearities in the receive chain or in the form of requiring discrete mixers coupled to the receive coils (49).

We hypothesize that a radar-based solution can potentially recover richer motion information than the conventional pilot tone by leveraging the exceptionally wide electromagnetic spectrum available for use inside the shielded MRI room. By contrast, other biomedical applications cannot use most of this spectrum due to regulatory restrictions. This potential is predicated on the fact that the propagation and scattering of electromagnetic waves depend on both the frequency and the dielectric properties of intervening tissues (50). As also exemplified in our results (Figure 4), this suggests that certain frequencies are more favorable than others for a given target structure. The frequency dependence of the motion signals observed in our phantom and respiratory experiments hints at the possibility of tailoring the radar system to a specific organ or motion type of interest, suggesting an avenue for future research. It is likely that the motion signals demonstrated in volunteer studies, employing CW radar at 1 and 4 GHz, predominantly represent reflections at the air-skin interface. Importantly, however, the phantom results demonstrate the radar’s capability for detecting internal motion. When tracking the internal mobile compartment, the motion signals improved at lower frequencies, consistent with the expected better penetration through the water-filled outer compartment. To unlock the full potential of radar for MRI motion correction, future work should exploit the wide frequency range in combination with advanced radar regimes such as FMCW or MIMO configurations, which provide richer information content than the comparatively simple CW radar system implemented in this pilot study. In this regard, the modular architecture of the software-defined radar platform enables experimentation with various radar architectures.

In a broader medical imaging context, the independence from the MR RF system opens up applications beyond MRI, as has been proposed for CT (51, 52) or for procedures in interventional radiology and radiation oncology.

While we successfully validated the radar phase shift against the image-based ground truth, a limitation of our study is the lack of a general analytical model for the observed signals. In this context, we refer the reader to a recent study by Lee et al. (20), which includes a comprehensive overview of CW radar theory as applied to MRI. The scenario in the MRI bore is different from most established radar applications insofar as the distance between the antennas and the subject is very short. It should be cautioned that depending on the frequency, the subject may be located in the near field, violating the assumptions of the classic Doppler equations. However, a quantitative signal model may not be required for all applications, for example, to reject motion-corrupted k-space lines or in situations where the motion model can be learned from an MR-based ground truth (23). Generating patient-specific motion models based on calibrating radar information with cine MRI represents an avenue for future research.

The antenna design was chosen for its ultra-wideband performance, enabling experiments over a broad frequency range. It should be noted, however, that this design was repurposed from a prior project, owing to the pilot character of this study. Dedicated antenna designs tailored for in-bore use can improve the electromagnetic compatibility with the MRI system, as described in the literature (53). While the antennas were mounted on a scaffold for the research prototype, a future transition to a clinically useful tool would require embedding the antennas under the bore liner. Tight space constraints and electromagnetic interactions with the MRI RF and gradient systems will require dedicated engineering efforts to balance MR compatibility and radar performance.

## Conclusion

We have developed an open-source, vendor-independent radar platform for MRI motion correction and demonstrated its effectiveness for a range of MRI techniques in both phantom and volunteer studies. The independence from the MRI scanner hardware, coupled with the flexibility of software-defined radio, offers broad potential for motion correction across imaging applications and body regions.

## Supporting information

Video S3: Illustrating the fidelity of the organ segmentation

Video S4: Multi-modal motion tracking in the head and neck.

## Data Availability Statement

Software and data used in the phantom experiments are currently being prepared for public release and will be available following formal acceptance of the manuscript. Due to regulatory restrictions, volunteer data cannot be shared publicly.

## Acknowledgments

We thank Sebastian Flassbeck and Ruoxun Zi for insightful discussions on MR instrumentation as well as Mahesh B. Keerthivasan and Gregor Körzdörfer for vendor support. Large language models were used to assist with the copyediting of this manuscript. The authors reviewed and edited all AI-generated suggestions and take full responsibility for the final content.

## Supporting Information

**Figure S1: Radar-based prospective motion management**.

**Figure S2: Investigating the signal origin of respiration-induced phase shift**.

**Video S3: Illustrating the fidelity of the organ segmentation**

**Video S4: Multi-modal motion tracking in the head and neck**.

## References

1. American College of Radiology, ACR Appropriateness Criteria. (2025). Available at: https://acsearch.acr.org/list [Accessed 3 July 2025].

2. World Health Organization, WHO list of priority medical devices for cancer management (2017).

3. J. B. Andre, et al., Toward quantifying the prevalence, severity, and cost associated with patient motion during clinical MR examinations. Journal of the American College of Radiology : JACR 12, 689–95 (2015).

4. A. van der Kouwe, J. B. Andre, Eds., Motion correction in MR: correction of position, motion, and dynamic changes (Academic Press is an imprint of Elsevier, 2023).

5. E. M. Schrauben, G. Lima da Cruz, C. W. Roy, T. Küstner, Motion Mitigation Techniques for Abdominal and Cardiac MR Imaging. J Magn Reson Imaging 63, 917–937 (2026).

6. Y. Wang, P. J. Rossman, R. C. Grimm, S. J. Riederer, R. L. Ehman, Navigator-echo-based real-time respiratory gating and triggering for reduction of respiration effects in three-dimensional coronary MR angiography. Radiology 198, 55–60 (1996).

7. A. C. S. Brau, J. H. Brittain, Generalized self-navigated motion detection technique: Preliminary investigation in abdominal imaging. Magn Reson Med 55, 263–270 (2006).

8. T. Niendorf, L. Winter, T. Frauenrath, “Electrocardiogram in an MRI Environment: Clinical Needs, Practical Considerations, Safety Implications, Technical Solutions and Future Directions” in Advances in Electrocardiograms-Methods and Analysis, R. Millis, Ed. (InTech, 2012).

9. D. W. Chakeres, A. Kangarlu, H. Boudoulas, D. C. Young, Effect of static magnetic field exposure of up to 8 Tesla on sequential human vital sign measurements. J Magn Reson Imaging 18, 346–352 (2003).

10. B. Madore, et al., External Hardware and Sensors, for Improved MRI. Magnetic Resonance Imaging 57, 690–705 (2023).

11. F. Preiswerk, et al., Hybrid MRI-Ultrasound acquisitions, and scannerless real-time imaging. Magn Reson Med 78, 897–908 (2017).

12. L. M. Gottwald, et al., Retrospective Camera-Based Respiratory Gating in Clinical Whole-Heart 4D Flow MRI. J Magn Reson Imaging 54, 440–451 (2021).

13. A. Andreychenko, et al., Thermal noise variance of a receive radiofrequency coil as a respiratory motion sensor. Magn Reson Med 77, 221–228 (2017).

14. A. T. Hess, E. M. Tunnicliffe, C. T. Rodgers, M. D. Robson, Diaphragm position can be accurately estimated from the scattering of a parallel transmit RF coil at 7 T. Magn Reson Med 79, 2164–2169 (2018).

15. D. Buikman, T. Helzel, P. Röschmann, The rf coil as a sensitive motion detector for magnetic resonance imaging. Magnetic resonance imaging 6, 281–9 (1988).

16. V. M. Runge, J. K. Richter, J. T. Heverhagen, Motion in Magnetic Resonance: New Paradigms for Improved Clinical Diagnosis. Invest Radiol 54, 383–395 (2019).

17. P. Speier, M. Fenchel, R. Rehner, PT-Nav: a novel respiratory navigation method for continuous acquisitions based on modulation of a pilot tone in the MR-receiver in ESMRMB, (2015).

18. E. Solomon, et al., Free-breathing radial imaging using a pilot-tone radiofrequency transmitter for detection of respiratory motion. Magnetic resonance in medicine 85, 2672–2685 (2021).

19. S. Anand, M. Lustig, Beat Pilot Tone (BPT): Simultaneous MRI and RF motion sensing at arbitrary frequencies. Magn Reson Med 92, 1768–1787 (2024).

20. W. Lee, et al., MRI retrospective respiratory gating and cardiac sensing by CW Doppler radar: A feasibility study. IEEE Trans Biomed Eng PP (2024).

21. J. Diao, et al., Frequency Modulated Continuous Wave Radar-based respiratory motion correction for cardiac MRI: Initial Results in Proc. Intl. Soc. Mag. Reson. Med., (2024).

22. F. Thiel, M. Hein, U. Schwarz, J. Sachs, F. Seifert, Combining magnetic resonance imaging and ultrawideband radar: a new concept for multimodal biomedical imaging. Rev Sci Instrum 80, 014302 (2009).

23. T. Neumann, et al., Ultra-wide band radar for prospective respiratory motion correction in the liver. Phys Med Biol 68 (2023).

24. C. G. Caro, J. A. Bloice, Contactless apnoea detector based on radar. Lancet 2, 959–961 (1971).

25. S. Dong, et al., A Review on Recent Advancements of Biomedical Radar for Clinical Applications. IEEE Open J. Eng. Med. Biol. 5, 707–724 (2024).

26. R. G. Machado, A. M. Wyglinski, Software-Defined Radio: Bridging the Analog–Digital Divide. Proc. IEEE 103, 409–423 (2015).

27. E. Yavari, O. Boric-Lubecke, S. Yamada, “Radar Principles” in Doppler Radar Physiological Sensing, (John Wiley & Sons, Ltd, 2016), pp. 21–38.

28. E. Hedayati, et al., An experimental system for detection and localization of hemorrhage using ultra-wideband microwaves with deep learning. Commun Eng 3, 126 (2024).

29. B. M. Schaller, A. W. Magill, R. Gruetter, Common Modes & Cable Traps in Proc. Intl. Soc. Mag. Reson. Med., (2011).

30. F. Preiswerk, et al., RF-sensing for trigger-based synchronization of auxiliary devices, and pulse-sequence debugging in Proc. Intl. Soc. Mag. Reson. Med., (2017), p. 4444.

31. J. Long, et al., GNU Radio. (2024). 10.5281/ZENODO.11042907. Deposited 22 April 2024.

32. I. Lenz, J. Holtom, A. Herschfelt, Y. Rong, D. Bliss, Respiratory and Heart Rate Detection Using Continuous-Wave Radar Testbed Implemented in GNU Radio. Proceedings of the GNU Radio Conference 7 (2022).

33. J. Demel, et al., Vector-Optimized Library of Kernels (VOLK). (2025). 10.5281/ZENODO.3360942. Deposited 3 February 2025.

34. N. B. Truong, Y.-J. Suh, C. Yu, Latency Analysis in GNU Radio/USRP-Based Software Radio Platforms in MILCOM 2013 - 2013 IEEE Military Communications Conference, (IEEE, 2013), pp. 305–310.

35. The GNU Radio Foundation, Inc., The signal metadata format (SigMF). (2018).

36. Abraham. Savitzky, M. J. E. Golay, Smoothing and Differentiation of Data by Simplified Least Squares Procedures. Anal. Chem. 36, 1627–1639 (1964).

37. E. Solomon, L. Alon, An open-source platform for simulating internal organ motion: Preliminary results in Proc. Intl. Soc. Mag. Reson. Med., (2023).

38. S. Zhang, K. T. Block, J. Frahm, Magnetic resonance imaging in real time: advances using radial FLASH. J Magn Reson Imaging 31, 101–109 (2010).

39. L. Feng, et al., XD-GRASP: Golden-angle radial MRI with reconstruction of extra motion-state dimensions using compressed sensing. Magn Reson Med 75, 775–788 (2016).

40. M. Uecker, et al., Berkeley advanced reconstruction toolbox in Proc. Intl. Soc. Mag. Reson. Med., (2015), p. 2486.

41. K. T. Block, M. Uecker, J. Frahm, Undersampled radial MRI with multiple coils. Iterative image reconstruction using a total variation constraint. Magnetic Resonance in Medicine 57, 1086–1098 (2007).

42. L. Feng, et al., Golden-angle radial sparse parallel MRI: Combination of compressed sensing, parallel imaging, and golden-angle radial sampling for fast and flexible dynamic volumetric MRI. Magnetic Resonance in Medicine 72, 707–717 (2014).

43. B. C. Lowekamp, D. T. Chen, L. Ibáñez, D. Blezek, The Design of SimpleITK. Front Neuroinform 7, 45 (2013).

44. T. Kober, J. P. Marques, R. Gruetter, G. Krueger, Head motion detection using FID navigators. Magn Reson Med 66, 135–143 (2011).

45. R. Frost, et al., Navigator-based reacquisition and estimation of motion-corrupted data: Application to multi-echo spin echo for carotid wall MRI. Magn Reson Med 83, 2026–2041 (2020).

46. N. Ravi, et al., SAM 2: Segment Anything in Images and Videos. [Preprint] (2024). Available at: http://arxiv.org/abs/2408.00714 [Accessed 11 June 2025].

47. S. M. Pizer, R. E. Johnston, J. P. Ericksen, B. C. Yankaskas, K. E. Muller, Contrast-limited adaptive histogram equalization: speed and effectiveness in [1990] Proceedings of the First Conference on Visualization in Biomedical Computing, (IEEE Comput. Soc. Press, 1990), pp. 337–345.

48. R. Zi, et al., Correction of Respiratory Motion in Free-Breathing DCE-MRI Using a Pilot-Tone Coil. NMR Biomed 39, e70278 (2026).

49. J. Kang, S. Anand, M. Lustig, External RF mixer for platform-independent motion sensing with beat pilot tone in Proc. Intl. Soc. Mag. Reson. Med., (2025), p. 5003.

50. C. Gabriel, “Compilation of the Dielectric Properties of Body Tissues at RF and Microwave Frequencies“ (Defense Technical Information Center, 1996).

51. F. Pfanner, J. Maier, T. Allmendinger, T. Flohr, M. Kachelrieß, Monitoring internal organ motion with continuous wave radar in CT. Med Phys 40, 091915 (2013).

52. A. Schwarz, et al., Free-Breathing Respiratory Triggered High-Pitch Lung CT: Insights From Phantom and Patient Scans. Invest Radiol 60, 517–525 (2025).

53. U. Schwarz, F. Thiel, F. Seifert, R. Stephan, M. A. Hein, Ultrawideband Antennas for Magnetic Resonance Imaging Navigator Techniques. IEEE Trans. Antennas Propagat. 58, 2107–2112 (2010).

