## Supplementary figures for "Software-defined Radar for MRI Motion Correction: A versatile, vendor-independent Platform"

### Supplementary material

Christoph Maier[1,2,†], Eddy Solomon[3], George Verghese[1,2], Hersh Chandarana[1,2], Kai Tobias Block[1,2*], and Leeor Alon[1,2*]

1: Bernard and Irene Schwartz Center for Biomedical Imaging, Department of Radiology, New York University Grossman School of Medicine, New York, NY, United States

2: Center for Advanced Imaging Innovation and Research (CAI2R), Department of Radiology, New York University Grossman School of Medicine, New York, NY, United States

3: Department of Biomedical Engineering, Technion – Israel Institute of Technology: Haifa, Israel

*: K.T.B. and L.A. contributed equally to this work

†: Corresponding author

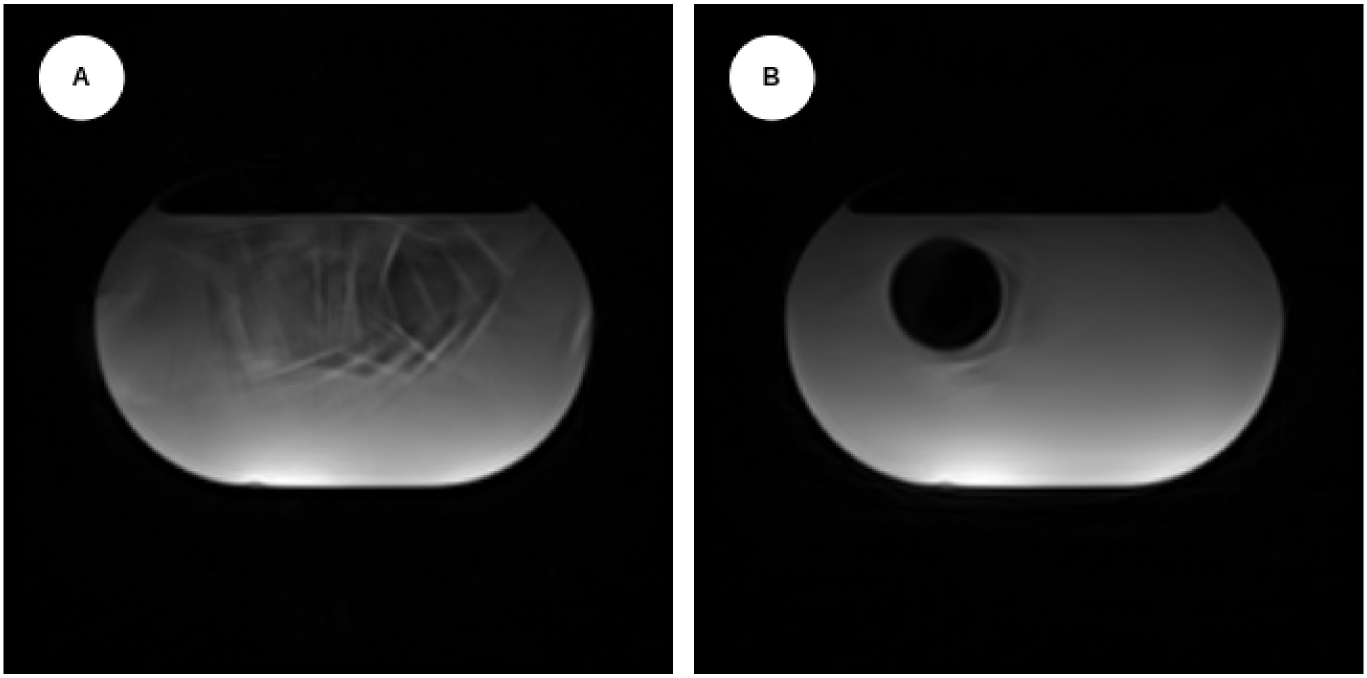


Figure 1: **Figure S1: Radar-based prospective motion management.**
(A) Untriggered T2w BLADE acquisition, with the inner compartment undergoing a periodic motion pattern. (B) Separate T2w BLADE acquisitions of the phantom undergoing the same motion pattern, with the SDR feeding trigger signals to the scanner based on the radar phase shift for prospective motion triggering. By only acquiring k-space data in a defined motion state, this approach effectively suppresses motion artifacts. Note: The inner compartment appears hypointense on T2w images due to the T2 shortening effect of gadolinium.

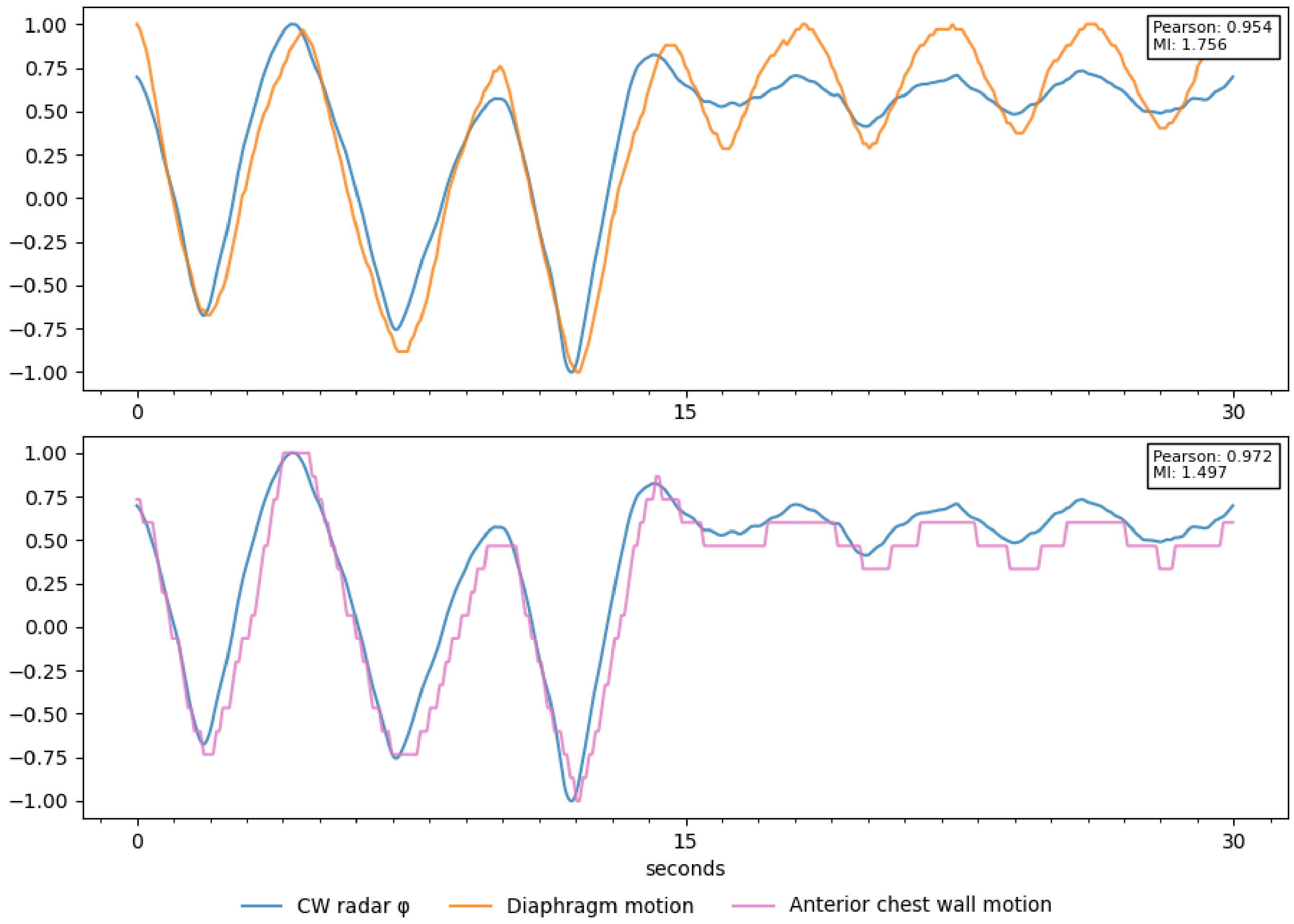


Figure 2: **Figure S2: Investigating the signal origin of respiration-induced phase shift.** A volunteer was instructed to perform deep breathing followed by shallow breathing. The CW Doppler radar phase shift (at a carrier frequency of 1 GHz) is compared to the head-foot movement of the diaphragm and to the anteroposterior motion of the anterior chest wall, as derived from spoiled GRE cine MRI at 11.4 fps. While the periodicity of the radar-based motion signal is in excellent agreement with both anatomical references, its amplitude over the entire dynamic range correlates better with chest wall displacement than with diaphragm motion. This suggests that reflection at the air-skin interface is the dominant source of the radar signal under these operating conditions, consistent with electromagnetic theory. Note that all signals have been normalized for visualization; the discretized appearance and lower mutual information of the chest wall motion is due to the low absolute amplitude of the chest wall excursions of a few millimeters.

**Video S3: Illustrating the fidelity of the organ segmentation**

**Video S4: Multi-modal motion tracking in the head and neck.**
